# Incidence and Outcomes of Atrial Arrhythmia with CDK4/6 Inhibitors in HR-Positive / HER2-Negative Breast Cancer

**DOI:** 10.1101/2025.08.01.25332600

**Authors:** Nathaniel E. Davis, Joerg Herrmann, David O. Hodge, Sophia Blumenfeld, Kathryn J. Ruddy, Nicholas Y. Tan

**Affiliations:** Department of Internal Medicine, Mayo Clinic, Rochester, MN, USA; Department of Cardiovascular Medicine, Mayo Clinic, Rochester, MN, USA; Department of Oncology, Mayo Clinic, Rochester, MN, USA; Department of Quantitative Health Sciences, Mayo Clinic, Jacksonville, FL, USA

## Abstract

**Background:** Cyclin-dependent kinase 4/6 (CDK4/6) inhibitors are increasingly used in hormone receptor-positive, HER2-negative (HR+/HER2−) breast cancer, yet emerging data suggest potential cardiotoxicity, including atrial arrhythmias (AA). Understanding the incidence and outcomes of AA in this population is essential as indications for CDK4/6 inhibitors expand.

**Objectives:** To evaluate the incidence of new-onset AA and associated outcomes in patients with HR+/HER2− breast cancer treated with CDK4/6 inhibitors.

**Methods:** We conducted a retrospective cohort study of patients treated at Mayo Clinic from 2015– 2024 who received CDK4/6 inhibitors for HR+/HER2− breast cancer. The primary outcome was incidence of AA (atrial fibrillation, atrial flutter, or atrial tachycardia). Secondary outcomes included cerebrovascular events and all-cause mortality. Kaplan-Meier estimates and Cox regression models were used to assess outcomes and associated risk factors.

**Results:** Among 2,782 patients, 59% received palbociclib, 28% abemaciclib, and 14% ribociclib.

New-onset AA occurred in 45, with cumulative incidence at 5 years of 2.8%. No significant differences in AA incidence were observed between agents (p=0.44). Multivariate analysis identified age at treatment as the only independent predictor of AA (HR 1.073, p<0.001). Four patients with new-onset AA experienced cerebrovascular events. New-onset AA was associated with increased mortality (HR 1.56, p=0.012).

**Conclusions:** CDK4/6 inhibitor therapy was associated with a low but clinically significant risk of new-onset AA, which in turn is associated with increased mortality. There was no significant difference in new-onset AA risk between different individual CDK4/6 inhibitor agents.

Prospective studies are needed to define mechanisms and guide monitoring strategies.

## Introduction

Historically, endocrine therapy has been the mainstay treatment for metastatic hormone receptor-positive / human epidermal growth factor receptor-negative (HR+/HER2-) breast cancer. The cyclin-dependent kinase 4/6 (CDK4/6) inhibitors– palbociclib, ribociclib, and abemaciclib – have been shown to have a synergistic effect in combination with ET. ^1–3^ Furthermore, CDK4/6 inhibitors are now included in adjuvant therapy for many patients with stage 2-3 cancers.^4,5^

However, CDK 4/6 inhibitors may cause cardiovascular toxicities.^6–8^ Ribociclib, in particular, has been associated with QT interval prolongation, prompting guidelines for electrocardiographic (ECG) monitoring during initiation.^9,10^ More broadly, emerging data has implicated CDK4/6 inhibitors with an elevated risk of major adverse cardiovascular events including atrial arrhythmias (AA) and heart failure, which in turn may be associated with increased mortality.^7^ The evolving utilization of CDK4/6 inhibitors and expansion beyond patients with metastatic disease has increased the importance of accurate understanding of cardiotoxicity profiles of these medications. Therefore, we sought to characterize the incidence and outcomes of AA among patients with HR+/HER2- breast cancer who were treated with CDK4/6 inhibitors at our institution.

## Methods

### Study Population and Data Collection

The data that support the findings of this study are available from the corresponding author upon reasonable request. The study was approved by the Mayo Clinic Institutional Review Board (IRB). Only patients who had previously authorized consent to use their records for research purposes were included. Patients were included in the study if they were ≥ 18 years of age, received care at Mayo Clinic between 2015 and 2024, were prescribed CDK4/6 inhibitors, and had a diagnosis of HR+, HER-2 negative breast cancer. Those patients who had no confirmation of HR+/HER-2 negative status were excluded from the study. Those patients who did not provide research authorization were also excluded from the study.

### Baseline Characteristics and Outcomes

Baseline characteristics collected for the patient cohort included demographic variables (age at CDK4/6 inhibitor therapy, sex), comprehensive cardiac and vascular histories, and oncologic details (Table 1). Oncologic specific variables included age at cancer diagnosis, and age at first administered CDK4/6 inhibitor. Primary outcomes included atrial fibrillation, atrial tachycardia, and atrial flutter, collectively defined as AA. Patients with AA were identified and verified via manual chart review. Secondary outcomes included cerebrovascular accidents (CVA) and mortality.

**Table 1:**
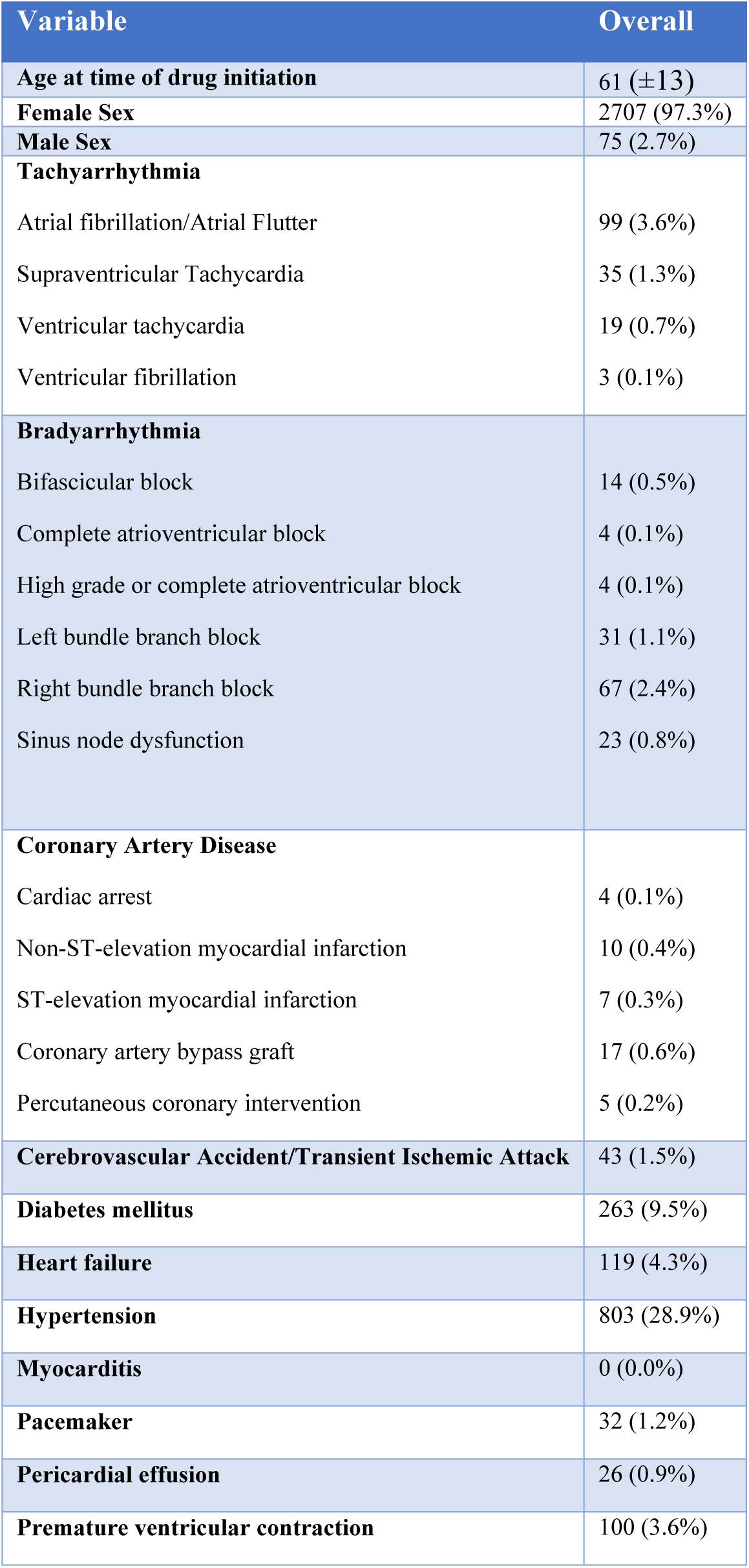
baseline cardiac and related conditions.

## Statistical Analysis

Baseline characteristics were analyzed using descriptive statistics (continuous variables – mean ±SD). Kaplan-Meier (KM) estimates were conducted for cumulative incidence of AA. This was done on all patients who received CDK 4/6 inhibitors, and for palbociclib, ribociclib, and abemaciclib independently. Univariate cox proportional hazards modeling with time to AA as the outcome of interest was performed, using age at treatment, sex, hypertension, diabetes, coronary artery disease, peripheral vascular disease, and stroke histories as variables of interest. Multivariate cox proportional hazards modeling was then performed, with age at treatment, sex, hypertension, coronary artery disease, and peripheral vascular disease included. KM curves were also created for all-cause mortality following CDK4/6 inhibitor drug initiation. Additionally, cox multivariate regression for mortality was performed with age at treatment, sex, and new-onset AA included.

## Results

### Baseline Characteristics and Treatment

A total of 2782 patients with a diagnosis of HR+/HER2- breast cancer were included in the study. Among these patients, 1630 (59%) patients were treated with palbociclib, 768 (28%) were treated with abemaciclib, and 384 (14%) were treated with ribociclib. The mean age at receipt of CDK4/6 inhibitor therapy was 61 (±13) years, and 2707 (97%) were female. The baseline cardiac and related conditions for the patient cohort are summarized in Table 1. Notable findings included a total of 803 (28.9%) patients with hypertension, 263 (9.5%) patients with diabetes mellitus, and 38 (1.4%) with ischemic heart disease (defined as cardiac arrest, myocardial infarction, Non-ST-elevation myocardial infarction, or ST-elevation myocardial infarction). 299 (10.7%) of patients had a history of arrhythmia. More specifically, 99 (3.6%) patients had a history of AA diagnosis prior to the initiation of CDK4/6 inhibitor therapy.

### Cumulative Incidence of AA

The 1-5-year cumulative incidences of new-onset AA were 1.1%, 1.5%, 2.3%, 2.6%, and 2.8% respectively (Figure 1A). There was no statistically significant difference between the three agents (p=0.44). KM curves for AA by drug are shown below (Figure 1B). One and five-year cumulative incidences of AA for abemaciclib ranged from 1.1% to 1.7%. Ribociclib had one-and five-year cumulative incidences of AA of 0.6% and 1% respectively. Palbociclib had the highest cumulative incidence of AA at 1.2% to 3.2% at 1 and 5 years, respectively. Detailed cumulative incidences are shown in Table 2.

**Figure 1:**
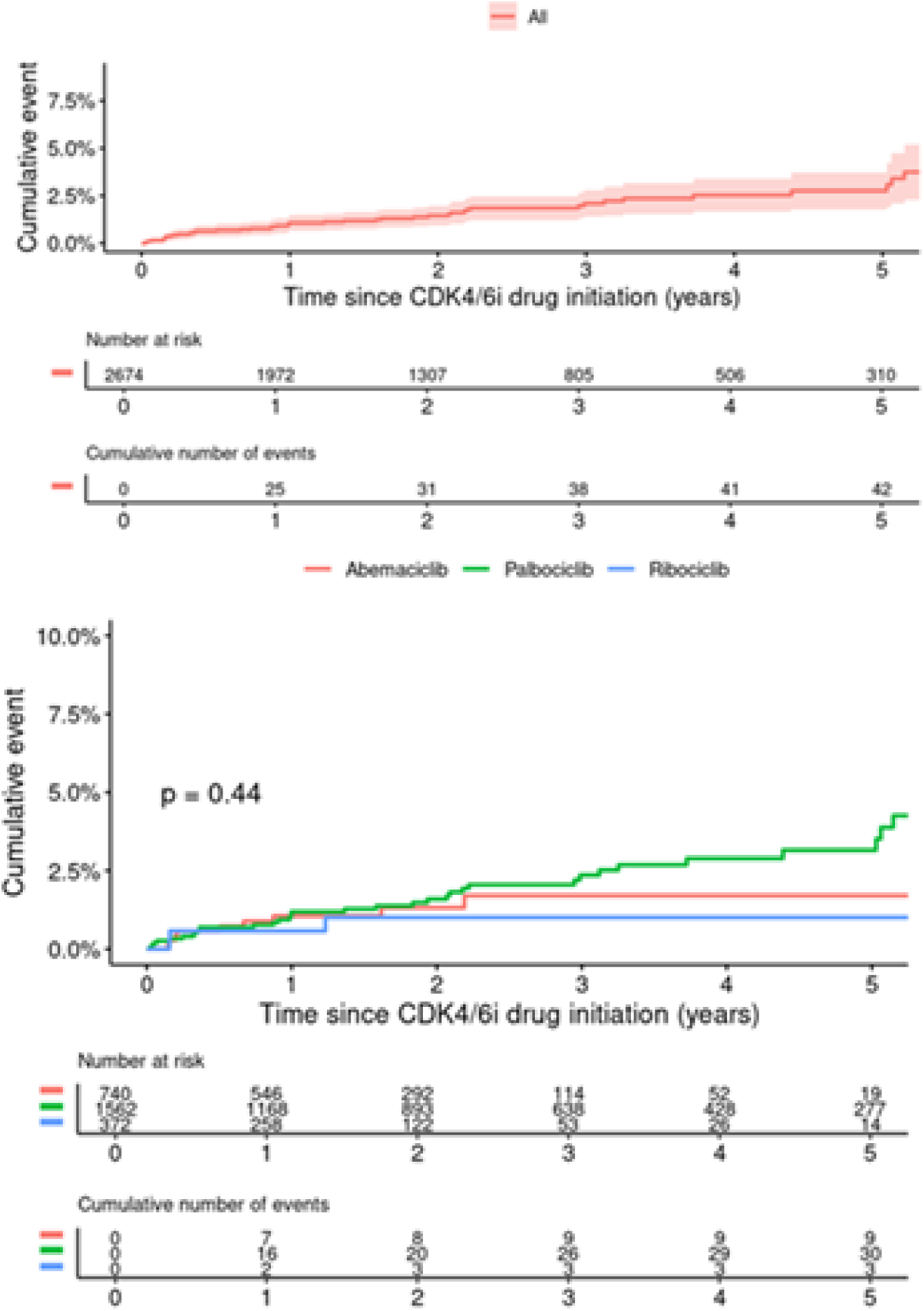
KM Curves for Incidence of AA. **Figure 1A**: KM curve for incidence of AA following initiation of CDK4/6 inhibitor therapy **Figure 1B**: KM curve for incidence of AA following initiation of CDK4/6 inhibitor therapy stratified by agent

**Table 2:** One to five-year cumulative incidences of AA stratified by CDK 4/6 inhibitor. 95% confidence intervals for estimates are provided.

| *Characteristic* | *1 Year* | *2 Year* | *3 Year* | *4 Year* | *5 Year* |
| --- | --- | --- | --- | --- | --- |
| *Abemaciclib* | *1.1% (0.3%, 1.8%)* | *1.3% (0.4%, 2.3%)* | *1.7% (0.5%, 2.9%)* | *1.7% (0.5%, 2.9%)* | *1.7% (0.5%, 2.9%)* |
| *Palbociclib* | *1.2% (0.6%, 1.8%)* | *1.6% (0.9%, 2.3%)* | *2.4% (1.4%, 3.3%)* | *2.9% (1.8%, 4.0%)* | *3.2% (1.9%, 4.4%)* |
| *Ribociclib* | *0.6% (0%, 1.4%)* | *1.1% (0%, 2.2%)* | *1.1% (0%, 2.2%)* | *1.1% (0%, 2.2%)* | *1.1% (0%, 2.2%)* |
| *Any Therapy* | *1.1% (0.7%, 1.6%)* | *1.5% (0.9%, 2.0%)* | *2.3% (1.5%, 3.0%)* | *2.6% (1.7%, 3.4%)* | *2.8% (1.8%, 3.8%)* |

On univariate analysis for time to AA, age at treatment (p<0.001), hypertension (p=0.007), and vascular disease (p< 0.001) were associated with new-onset AA (Table 3). However, following multivariate adjustment, only age at treatment was statistically significant (p<0.001) (Table 4).

**Table 3:** Univariate Cox modeling for new-onset atrial arrhythmia following CDK4/6 inhibitor therapy.

| Variable<sub>Nmiss= 99</sub> | HR | CI.lower.<br>HR | CI.upper<br>.HR | p-value |
| --- | --- | --- | --- | --- |
| **Age at Treatment** | 1.083 | 1.055 | 1.112 | < 0.001 |
| **Sex (Male)** | 2.588 | 0.625 | 10.709 | 0.189 |
| **Hypertension (Yes)** | 2.197 | 1.245 | 3.877 | 0.007 |
| **Diabetes (Yes)** | 1.542 | 0.655 | 3.629 | 0.322 |
| **Vascular Disease (Yes)** | 3.589 | 1.986 | 6.485 | < 0.001 |
| **Stroke (Yes)** | 1.883 | 0.260 | 13.654 | 0.531 |

**Table 4:** Multivariate Cox modeling for new-onset atrial arrhythmia following CDK4/6 inhibitor therapy.

| Variable<sub>N<sub>miss</sub>= 99</sub> | HR | CI<br>lower | CI<br>upper | p-value |
| --- | --- | --- | --- | --- |
| Age at treatment | 1.073 | 1.043 | 1.105 | < 0.001 |
| Sex (Male) | 2.202 | 0.517 | 9.386 | 0.286 |
| Hypertension (Yes) | 1.076 | 0.567 | 2.043 | 0.823 |
| Vascular Disease (Yes) | 1.776 | 0.898 | 3.513 | 0.099 |
| Stroke (Yes) | 0.720 | 0.093 | 5.543 | 0.752 |

### Cerebrovascular Accidents and Mortality Following New-Onset Atrial Arrhythmias

Of the patients who developed AA after initiation of CDK4/6 inhibitor therapy, a total of 4 patients experienced CVA. All four patients experienced CVA within the first year of diagnosis with AA. 2 of the four patients were anticoagulated at the time of CVA. Of the two patients that were not anticoagulated, one was unable to receive therapeutic anticoagulation due to bleeding risk, and the other was diagnosed with AA at the time they experienced CVA. None of the patients had undergone left atrial appendage occlusion.

Mortality by any cause within five years after the initiation of CDK4/6 inhibitor therapy was also analyzed for our patient cohort. Mortality at 1 and 5 years were 12.4% and 41.2% respectively (Figure 2). After adjustment for age at treatment and sex, new onset AA was significantly associated with all-cause mortality (hazard ratio 1.56, 95% CI 1.102-2.202, p=0.012) (Table 5).

**Figure 2:**
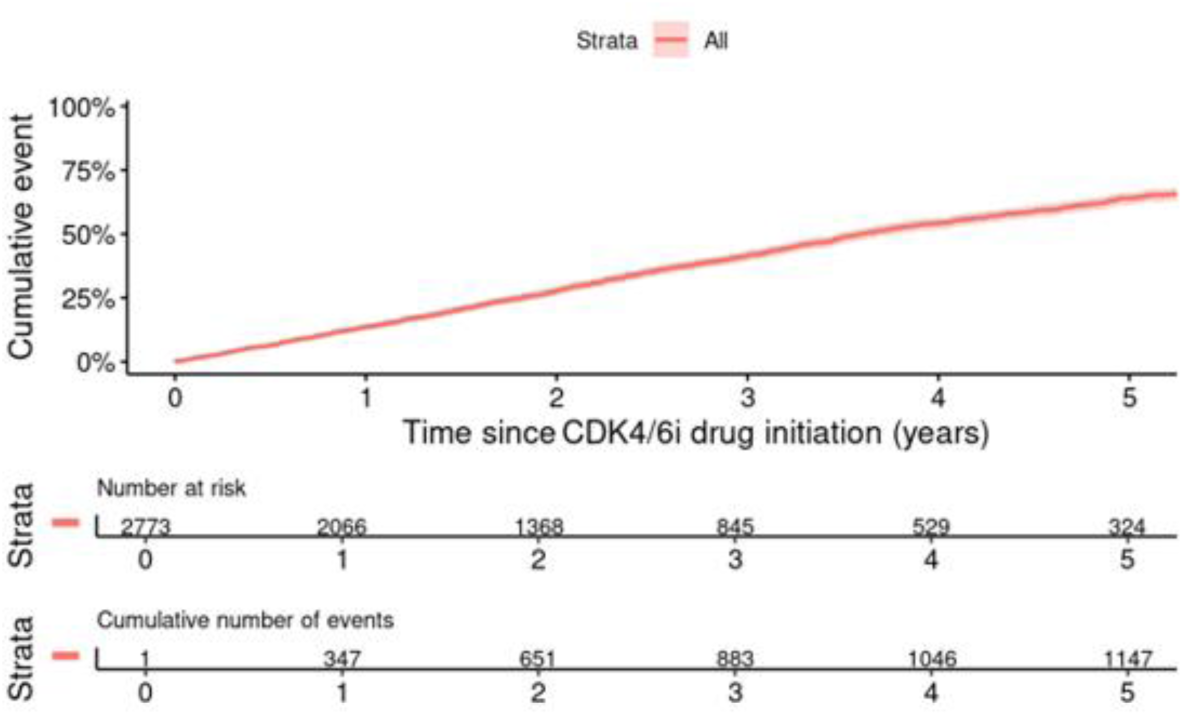
KM curve for all-cause mortality in patients with time since CDK4/6 inhibitor therapy on the x-axis and number at risk, with the cumulative number of deaths included below.

**Table 5:** Cox Multivariate Regression for Mortality.

| Variable | HR | CI lower | CI upper | p-value |
| --- | --- | --- | --- | --- |
| Age at treatment | 1.016 | 1.012 | 1.021 | < 0.001 |
| Sex (Male) | 2.494 | 1.884 | 3.302 | < 0.001 |
| AA | 1.558 | 1.102 | 2.202 | 0.012 |

## Discussion

In this study of 2782 patients with HR+/HER2-breast cancer who received CDK4/6 inhibitors, we sought to ascertain the incidence, risk factors, and outcomes of new-onset AA. The following salient findings were identified: 1. The 5-year risk of new-onset AA following CDK4/6 initiation was 2.8%; 2. There was no significant difference in AA cumulative incidence among the 3 CDK4/6 inhibitor agents; 3. Age was the only independent predictor for new-onset AA; 4. New-onset AA was associated with a 56% mortality risk increase after adjusting for age and sex. To our knowledge, this is the largest investigation of the potential link between CDK4/6 inhibitor use and AA development.

Cancer and heart disease share a bidirectional relationship that continues to evolve with advances in therapies across both disciplines. CDK4/6 inhibitors have significantly changed the management and natural history of HR+/HER2-breast cancer; however, as patients survive longer, potential cardiac adverse effects may become apparent. There are several possible explanations for new-onset AA in patients taking CDK 4/6 inhibitors including alteration in ion channel function, structural heart changes, and inflammatory and fibrotic changes in the myocardium.^7,12,13^ Reassuringly, the incidence of AA in our study is similar to that previously identified across a broader cohort of patients with breast cancer^14,15^, suggesting that CDK4/6 inhibitor may not excessively increase the risk of AA onset.

On univariate analysis, age at treatment, hypertension, and vascular disease, including coronary artery disease and peripheral artery disease, all were statistically significantly associated with new onset AA, but history of stroke, diabetes, and sex were not. However, on multivariate analysis, only age at treatment was statistically significant. The relevance of age suggests that older patients, particularly those with other risk factors for developing arrhythmia, should be more closely monitored for AA during therapy.

Of the patients that developed new onset AA after initiation of CDK4/6 inhibitor therapy, a total of four experienced CVA. Each of these events occurred within the first year of diagnosis with AA. It is widely recognized that new onset AA in patients with malignancy is associated with worse outcomes than in those patients without malignancy.^7,16,17^ Despite the relatively small cohort of patients with new onset AA, the associated stroke rate (9.5%) among those affected underscores the potential clinical impact of these arrhythmias.

All-cause mortality in patients treated with CDK4/6 inhibitors was 41.2% within five years of initiation of therapy. Cox multivariate regression models including age at treatment, sex, and new onset AA were statistically significantly associated with mortality. This is consistent with prior studies identifying AA as a risk factor for mortality in patients with cancer.^7^ The complex interplay between AA and cancer is well established. Inflammatory pathways activated in patients with cancer have been shown to contribute to atrial remodeling and increase susceptibility to arrhythmias.^18–20^ As such, the presence of AA may serve as a marker of systemic illness, heightened inflammatory states, and overall disease burden. Furthermore, AA may directly contribute to increased morbidity and mortality by adversely affecting quality of life, promoting the development or progression of heart failure, and inducing maladaptive cardiac remodeling.

## Limitations

This study is subject to the limitations inherent in retrospective designs, including potential misclassification and residual confounding. While all AA diagnoses were manually verified, reliance on ICD coding for baseline comorbidities and events introduces the possibility of underreporting or misdiagnosis. Furthermore, data on prior chemotherapy, radiation therapy, or surgical interventions were self-reported and may lack granularity. Variability in follow-up due to referral patterns may also have impacted outcome capture. By design, all patients in this study received CDK4/6 inhibitors. We elected not to perform a comparative analysis against a historical control group (when CDK4/6 inhibitors were not available) given expected major differences in clinical characteristics and outcomes. However, this limits our ability to tease out the effect of CDK4/6 inhibitor on new-onset AA independent of the impact of breast cancer or other aspects of breast cancer treatment.

## Conclusion

Patients who receive CDK4/6 inhibitors for breast cancer experience a low but measurable risk of new-onset atrial arrhythmias (AA). No significant differences in arrhythmia incidence were observed among the three CDK4/6 inhibitor agents. New-onset AA was associated with increased mortality in this cohort. Prospective studies are needed to clarify underlying mechanisms, improve risk stratification, and develop preventive strategies.

## Abbreviations

HR+: Hormone Receptor-Positive
HER2-: Human Epidermal Growth Factor Receptor 2-Negative
CDK4/6: Cyclin-Dependent Kinase 4/6
AA: Atrial Arrhythmia
ECG: Electrocardiogram
CVA: Cerebrovascular Accident
KM: Kaplan-Meier
HR: Hazard Ratio
CI: Confidence Interval
IRB: Institutional Review Board

## Data Availability

All data utilized in the manuscript is available upon request

## Acknowledgements

none

